# Kidney Biomarkers and Major Adverse Kidney Events in Critically Ill Patients

**DOI:** 10.1101/2020.06.06.20124354

**Authors:** Alexander H. Flannery, Katherine Bosler, Victor Ortiz-Soriano, Fabiola Gianella, Victor Prado, Robert D. Toto, Orson W. Moe, Javier A. Neyra

## Abstract

**Background:** Several biomarkers of acute kidney injury (AKI) have been examined for their ability to predict AKI earlier than serum creatinine. Few studies have focused on using kidney biomarkers to better predict major adverse kidney events (MAKE), an increasingly used composite outcome in critical care nephrology research.

**Methods:** Single-center prospective study collecting blood and urine samples from critically ill patients with AKI KDIGO stage 2 or above, and matched controls from a single, tertiary care intensive care unit. Samples were collected at 24-48 hours after AKI diagnosis (cases) or ICU admission (controls), 5-7 days later, and 4-6 weeks following discharge for AKI patients. The primary outcome of interest was MAKE at hospital discharge.

**Results:** Serum/urinary neutrophil gelatinase-associated lipocalin, serum/urinary cystatin C, and urinary kidney injury molecule-1 early in the AKI or ICU course were all significantly higher in patients with MAKE compared to those not experiencing MAKE at hospital discharge. Serum cystatin C, and to a lesser extent serum NGAL, significantly improved upon a clinical prediction model of MAKE as assessed by the area under the receiver operating characteristic curve.

Patients without MAKE experienced a greater decline in serum NGAL from initial measurement to second measurement than those patients experiencing MAKE.

**Conclusion:** Early measures of kidney biomarkers in critically ill patients are associated with MAKE. This relationship appears to be greatest with serum NGAL and cystatin C, which display additive utility to a clinical prediction model. Trending serum NGAL may also have utility in predicting MAKE.

## Introduction

The last decade has seen an explosion of research on biomarkers of acute kidney injury (AKI). Over 15 different biomarkers have been investigated, with a combination of markers receiving US Food and Drug Administration (FDA) approval in 2014 for detecting AKI within the ensuing 12 hours: the product of tissue inhibitor of metalloproteinase 2 TIMP-2 and insulin-like growth factor binding protein 7 [TIMP-2] × [IGFBP7].^1, 2^ The primary focus of these studies was the search for a biomarker that would detect AKI sooner than serum creatinine, allowing earlier recognition to improve care.^3, 4^ A secondary aim of this line of research has been to improve risk stratification and longer term prognostication with kidney biomarkers. This is particularly relevant for patients in the intensive care unit (ICU), where the incidence of AKI is high and conversations with family and decisions regarding supportive therapies such as renal replacement therapy (RRT) can be facilitated by improved prognostication of AKI that extends beyond the period of the acute illness.

Major adverse kidney events (MAKE), the composite of death, need of RRT, or worsened kidney function, is increasingly recognized as an important, patient-centered outcome for studies of AKI, particularly in critically ill patients.^5^ Previous studies of biomarkers for AKI have primarily limited evaluation of associations with MAKE out to 30 days, thus we sought to assess the relationship with AKI at further time intervals from AKI onset. Further, limited literature has been published to predict MAKE, specifically from clinical models of ICU patients, but preliminary data suggest simple models considering severity of illness provide good performance predicting MAKE out to 30 days.^6^ Accordingly, we sought to assess kidney biomarker relationships with MAKE at various time points out to one year of follow-up and test the utility of adding biomarker data to a clinical prediction model for MAKE by hospital discharge.

## Methods

### Study Design and Participants

This was a secondary analysis of the AKI biobank from the “Klotho and Acute Kidney Injury” (KLAKI) Study Group.^7^ In brief, this single center, prospective study enrolled adult patients ≥ 18 years old over a one year period admitted to the ICU with stage ≥ 2 AKI per Kidney Disease: Improving Global Outcomes (KDIGO) criteria and matched controls based on age, gender, and baseline estimated glomerular filtration rate (eGFR).^8^ Patients must have had a baseline eGFR ≥ 60 mL/min/1.73 m^2^. Patients were excluded if they had end-stage kidney disease (ESKD), a kidney or any other solid organ transplant, evidence of AKI prior to ICU admission, or uroepithelial tumors.^7^

### Data Collection and Laboratory Analysis

Blood and urine samples were taken from all patients at an initial time point (T1), 24-48 hours after AKI diagnosis (cases) or ICU admission (controls). A second set of samples (T2) was obtained 5-7 days following this initial collection. For AKI patients, a third set (T3) was collected 4-6 weeks following discharge if patients survived and followed up with clinic visit. Blood and urine samples were centrifuged at 1,000*g* at 4°C for 10 minutes, supernatant aliquoted into cryovials, and stored at -80°C until biomarker measurement. Serum neutrophil gelatinase-associated lipocalin (NGAL) and cystatin C and urinary NGAL, cystatin C, and kidney injury molecule-1 (KIM-1) were measured using Enzyme-Linked Immunosorbent Assay (ELISA) (BioPorto Diagnostics, Needham, MA; R&D Systems, Minneapolis, MN). Capillary electrophoresis was used to measure urinary creatinine and serum creatinine was obtained from routine clinical care.^9^ Baseline demographics, comorbidities, and clinical data associated with the ICU admission were collected, including the Acute Physiology and Chronic Health Evaluation (APACHE) II and Charlson Comorbidity Index (CCI).^10, 11^

### Outcomes and Statistical Analysis

The primary study objective was to compare biomarker measurements between patients with and without the outcome of MAKE at the following time points: hospital discharge (DC), 90 days (90), 6 months (6 mo), and 1 year (1 yr). At the specified time point, MAKE was defined as any of the following: death, RRT dependence, or decrease in eGFR to <75% of baseline.^5^ Secondarily, we sought to assess whether biomarker measurements at time T1 improved upon a clinical prediction model for MAKE-DC. In an exploratory analysis, we examined the trajectory of the biomarkers between times T1 and T2 (the Δ change in biomarker= value at time T2 – value at time T1) in those patients with and without MAKE-DC.

Urinary biomarkers were normalized to urinary creatinine (Ucr) in reporting. Categorical data are presented as proportions and continuous data as medians with interquartile ranges or means ± standard deviation depending on the distribution. Categorial variables were compared using a chi-square or Fisher’s exact test, as appropriate. Continuous variables were compared using an independent samples t-test or Mann Whitney-U test. A logistic regression model for MAKE-DC was generated using a simple clinical prediction model with two variables, APACHE II and CCI based on the number of MAKE-DC events in the dataset. Collinearity was assessed with the use of variance inflation factors. The added value of individual biomarkers from T1 was assessed with their addition to the model with the use of the area under the receiver operating characteristic curve (AUROC). Statistical significance was considered a two-sided p value < 0.05. Analyses were performed in Stata (Version 16.1; Stata Corp, College Station, TX) and R (R Foundation for Statistical Computing, Vienna, Austria).

## Results

The study included a total of 106 patients, 54 with AKI and 52 matched controls. Patient characteristics are shown in **Table 1**. Per the study design, AKI and controls were not different in age, sex, or baseline eGFR. A greater proportion of AKI patients were in the medical ICU compared to a greater proportion of control patients that were admitted to the surgical ICU. AKI patients tended to be more acutely ill overall, as evidenced by higher APACHE II and a greater need for vasopressors and mechanical ventilation. AKI patients also had more liver disease and a higher CCI compared to controls.

**Table 1.** Patient Characteristics.

| <u>Characteristics</u> | <u>AKI (n=54)</u> | <u>No AKI (n=52)</u> | <u>p-value</u> |
| --- | --- | --- | --- |
| Age (years) | 55.8 ± 16.7 | 58.0 ± 14.5 | 0.471 |
| Sex (% male) | 31(57.4%) | 29 (55.8%) | 0.865 |
| Race | White-34(63.0%) | White-40(76.9%) | 0.183 |
|  | Black-12(22.2%) | Black-5(9.6%) |  |
|  | Other-8(14.8%) | Other-7(13.5%) |  |
| ICU Type | SICU-9(16.7%) | SICU-18(34.6%) | 0.001 |
|  | MICU-26(48.2%) | MICU-6(11.5%) |  |
|  | CCU-10(18.5%) | CCU-15(28.9%) |  |
|  | Other ICU-9(16.7%) | Other ICU-13(25%) |  |
| Diabetes (%) | 17(31.5%) | 10(19.2%) | 0.148 |
| Hypertension (%) | 27(50.0%) | 29(55.8%) | 0.552 |
| Heart Failure (%) | 15(27.8%) | 17(32.7%) | 0.582 |
| Liver Disease (%) | 18(33.3%) | 6(11.5%) | 0.007 |
| Cancer (%) | 23(42.6%) | 23(45.1%) | 0.796 |
| Baseline Serum Creatinine (mg/dl) | 0.86 ± 0.26 | 0.82 ± 0.20 | 0.323 |
| Baseline eGFR (ml/min) | 88 (69.5-104) | 92 (78.3-103) | 0.399 |
| APACHE II | 20 (16-26) | 12 (9-19) | <0.001 |
| Number Pressors/inotropes | 1.5 (0-3) | 0 (0-1) | 0.001 |
| Mechanical Ventilation (%) | 34 (63.0%) | 20 (38.5%) | 0.012 |
| Charlson Comorbidity Index | 4 (2.8-7) | 2 (2-4) | <0.001 |

Biomarker values at time points T1, T2, and T3 are compared between AKI patients and controls in **Table 2**. Seven AKI patients were anuric and thus unable to contribute urine specimens. Death, discharge, or loss to follow up were other reasons for absence of a repeated measurement at the later time points of T2 and T3. Serum and urine biomarkers at all time points were higher, often by several orders of magnitude, in patients with AKI compared to controls, with the exception of urinary KIM-1/Ucr at time point T2. Of note, AKI patients with measurements at time point T3, several weeks following AKI, demonstrated sustained elevations in biomarkers, particularly serum NGAL and cystatin C, that were as high or higher than values in the control group at time T1.

**Table 2.** Kidney Biomarkers Stratified by Presence or Absence of AKI.

| Biomarker | n | AKI (n=54) | n | No AKI (n=52) | p-value |
| --- | --- | --- | --- | --- | --- |
| Serum NGAL Time 1 (ng/ml) | 54 | 487 (342-1317) | 52 | 163 (115-213) | <0.001 |
| Serum NGAL Time 2 (ng/ml) | 29 | 441 (290-705) | 7 | 153 (118-185) | 0.001 |
| Serum NGAL Time 3 (ng/ml) | 22 | 233 (178-360) |  |  |  |
| Urinary NGAL/Ucr Time 1 (ng/ml) | 47 | 1305 (304-6068) | 52 | 32 (9-85) | <0.001 |
| Urinary NGAL/Ucr Time 2 (ng/ml) | 26 | 791 (301-2436) | 7 | 62 (29-82) | 0.001 |
| Urinary NGAL/Ucr Time 3 (ng/ml) | 20 | 72 (30-498) |  |  |  |
| Serum Cystatin C Time 1 (ng/ml) | 54 | 2122 (1189-2978) | 52 | 712 (576-842) | <0.001 |
| Serum Cystatin C Time 2 (ng/ml) | 29 | 1506 (1111-2258) | 7 | 833 (623-974) | 0.001 |
| Serum Cystatin C Time 3 (ng/ml) | 21 | 1299 (908-2387) |  |  |  |
| Urinary Cystatin C/Ucr Time 1 (ng/ml) | 47 | 785 (125-1561) | 52 | 48 (9-166) | <0.001 |
| Urinary Cystatin C/Ucr Time 2 (ng/ml) | 26 | 652 (182-1462) | 7 | 63 (17-128) | 0.003 |
| Urinary Cystatin C/Ucr Time 3 (ng/ml) | 19 | 68 (44-325) |  |  |  |
| Urinary KIM-1/Ucr Time 1 (ng/ml) | 47 | 5.7 (3.3-13.9) | 52 | 2.3 (1.2-4.4) | <0.001 |
| Urinary KIM-1/Ucr Time 2 (ng/ml) | 47 | 5.2 (3.1-8.1) | 7 | 4.1 (0.9-5.4) | 0.308 |
| Urinary KIM-1/Ucr Time 3 (ng/ml) | 20 | 2.3 (1.7-4.3) |  |  |  |

Time 1: 24-48 hours after AKI diagnosis (cases) or ICU admission (controls)
Time 2: 5-7 days following Time 1 collection
Time 3: 4-6 weeks following discharge if patient survived and followed up with clinic visit

All biomarkers assessed at time T1 were significantly more elevated in patients experiencing MAKE-DC than those not (**Table 3**). Serum NGAL and cystatin C were the only biomarkers at time T2 that were significantly higher in patients experiencing MAKE-DC compared to those not. These biomarker values at T1 and T2 between patients experiencing MAKE-DC and those not can be visualized in **Figure 1**. Serum/urinary NGAL and serum cystatin C measurements at time T1 were significantly higher in those patients with MAKE-DC, MAKE-90, MAKE-6 mo, and MAKE-1 yr. Urinary cystatin C/Ucr and urinary KIM-1/Ucr measurements at timepoint T1 were significantly higher in MAKE patients than non-MAKE patients out to six months, but not at 1 year.

**Table 3.** Kidney Biomarkers and Major Adverse Kidney Events.

| **Biomarker** | **MAKE at Discharge**<br>**(n=49)** | **No MAKE at**<br>**Discharge (n=57)** | **p-**<br>**value** | **MAKE at 90 Days**<br>**(n=47)** | **No MAKE at 90**<br>**Days (n=35)** | **p-**<br>**value** |
| --- | --- | --- | --- | --- | --- | --- |
| Serum NGAL Time 1 (ng/ml) | 485 (340-1190) | 175 (117-235) | <0.001 | 483 (244-1307) | 189 (119-304) | <0.001 |
| Serum NGAL Time 2 (ng/ml) | 452 (277-952) (n=23) | 163 (120-439) (n=13) | 0.011 | 441 (238-804) (n=21) | 157 (133-437) (n=7) | 0.062 |
| Serum NGAL Time 3 (ng/ml) |  |  |  | 237 (200-886) (n=12) | 238 (106-516) (n=5) | 0.574 |
| Urinary NGAL/Ucr Time 1 (ng/ml) | 1037 (283-6085) (n=42) | 38 (10-103) | <0.001 | 850 (140-3649) (n=40) | 34 (10-99) | <0.001 |
| Urinary NGAL/Ucr Time 2 (ng/ml) | 553 (82-2488) (n=20) | 109 (49-1534) (n=13) | 0.316 | 553 (75-1322) (n=18) | 109 (35-2496) (n=7) | 0.790 |
| Urinary NGAL/Ucr Time 3 (ng/ml) |  |  |  | 259 (40-891) (n=10) | 30 (15-561) (n=5) | 0.165 |
| Serum Cystatin C Time 1 (ng/ml) | 2283 (1153-3038) | 716 (579-923) | <0.001 | 2018 (837-3002) | 713 (574-1024) | <0.001 |
| Serum Cystatin C Time 2 (ng/ml) | 1565 (1117-2464) (n=23) | 974 (679-1424) (n=13) | 0.006 | 1369 (1100-2554) (n=21) | 962 (667-1403) (n=7) | 0.036 |
| Serum Cystatin C Time 3 (ng/ml) |  |  |  | 1668 (1201-2595) (n=12) | 1445 (813-3124) (n=4) | 0.599 |
| Urinary Cystatin C/Ucr Time 1 (ng/ml) | 611 (122-1601) (n=42) | 56 (14-620) | <0.001 | 339 (48-1529) (n=40) | 68 (17-837) | 0.047 |
| Urinary Cystatin C/Ucr Time 2 (ng/ml) | 593 (41-1616) (n=20) | 200 (92-726) (n=13) | 0.434 | 370 (75-1462) (n=18) | 147 (62-776) (n=7) | 0.574 |
| Urinary Cystatin C/Ucr Time 3 (ng/ml) |  |  |  | 69 (46-820) (n=9) | 116 (55-319) (n=5) | 0.947 |
| Urinary KIM-1/Ucr Time 1 (ng/ml) | 5.6 (3.0-13.4) (n=42) | 2.4 (1.2-6.1) | <0.001 | 5.2 (2.5-13.3) (n=40) | 2.3 (1.4-6.1) | 0.014 |
| Urinary KIM-1/Ucr Time 2 (ng/ml) | 5.2 (3.0-7.9) (n=20) | 4.1 (2.2-7.5) (n=13) | 0.650 | 4.6 (2.8-7.7) (n=18) | 5.2 (1.2-6.8) (n=7) | 0.701 |
| Urinary KIM-1/Ucr Time 3 (ng/ml) |  |  |  | 2.4 (1.8-4.9) (n=10) | 3 (1.9-4.3) (n=5) | 0.953 |
| **Biomarker** | **MAKE at 6 months**<br>**(n=45)** | **No MAKE at 6**<br>**months (n=34)** | **p-**<br>**value** | **MAKE at 1 year**<br>**(n=48)** | **No MAKE at 1 year**<br>**(n=27)** | **p-**<br>**value** |
| Serum NGAL Time 1 (ng/ml) | 474 (229-1203) | 193 (129-312) | <0.001 | 426 (188-927) | 192 (133-279) | 0 |
| Serum NGAL Time 2 (ng/ml) | 452 (272-1018) (n=17) | 158 (114-304) (n=8) | 0.011 | 321 (168-575) (n=20) | 153 (133-336) (n=3) | 0.31 |
| Serum NGAL Time 3 (ng/ml) | 328 (226-1277) (n=9) | 150 (108-238) (n=7) | 0.023 | 226 (177-1158) (n=10) | 213 (136-306) (n=4) | 0.64 |
| Urinary NGAL/Ucr Time 1 (ng/ml) | 510 (107-3277) (n=39) | 33 (9-150) (n=33) | <0.001 | 510 (92-3079) (n=41) | 33 (10-61) (n=27) | <0.001 |
| Urinary NGAL/Ucr Time 2 (ng/ml) | 494 (66-2009) (n=14) | 343 (46-1126) (n=8) | 0.664 | 459 (62-1832) (n=17) | 109 (35-1173) (n=3) | 0.69 |
| Urinary NGAL/Ucr Time 3 (ng/ml) | 416 (23-904) (n=7) | 42 (19-70) (n=7) | 0.209 | 259 (34-900) (n=8) | 43 (14-167) (n=4) | 0.15 |
| Serum Cystatin C Time 1 (ng/ml) | 2018 (830-3038) | 710 (571-999) | <0.001 | 1551 (801-2804) | 713 (560-989) | <0.001 |
| Serum Cystatin C Time 2 (ng/ml) | 1506 (1042-2677) (n=17) | 1035 (709-1340) (n=8) | 0.043 | 1358 (935-2598) (n=20) | 974 (833-1318) (n=3) | 0.31 |
| Serum Cystatin C Time 3 (ng/ml) | 2380 (1088-2708) (n=9) | 1205 (955-1479) (n=6) | 0.088 | 2206 (1097-2685) (n=10) | 940 (810-1766) (n=4) | 0.11 |
| Urinary Cystatin C/Ucr Time 1 (ng/ml) | 398 (48-1561) (n=39) | 57 (13-650) (n=33) | 0.036 | 280 (27-1299) (n=41) | 53 (11-643) | 0.11 |
| Urinary Cystatin C/Ucr Time 2 (ng/ml) | 503 (23-1462) (n=14) | 174 (79-568) (n=8) | 0.764 | 247 (23-782) (n=17) | 147 (63-1794) (n=3) | 0.84 |
| Urinary Cystatin C/Ucr Time 3 (ng/ml) | 437 (49-1477) (n=6) | 68 (49-116) (n=7) | 0.181 | 317 (69-1084) (n=7) | 52 (49-56) (n=4) | 0.07 |
| Urinary KIM-1/Ucr Time 1 (ng/ml) | 5.1 (2.5-13.3) (n=39) | 2.1 (1.1-5.8) (n=33) | 0.005 | 4.3 (1.6-13.2) (n=41) | 2.1 (1.4-5.5) | 0.09 |
| Urinary KIM-1/Ucr Time 2 (ng/ml) | 5.8 (2.8-7.7) (n=14) | 4.3 (1.6-6.4) (n=8) | 0.525 | 4.5 (2.1-7.3) (n=17) | 4.1 (3.7-5.2) (n=3) | 0.96 |
| Urinary KIM-1/Ucr Time 3 (ng/ml) | 4.5 (2.0-6.4) (n=7) | 2.3 (1.8-3.8) (n=7) | 0.209 | 2.2 (1.7-6.0) (n=8) | 2.9 (1.0-4.5) (n=4) | 0.93 |

**Figure 1.**
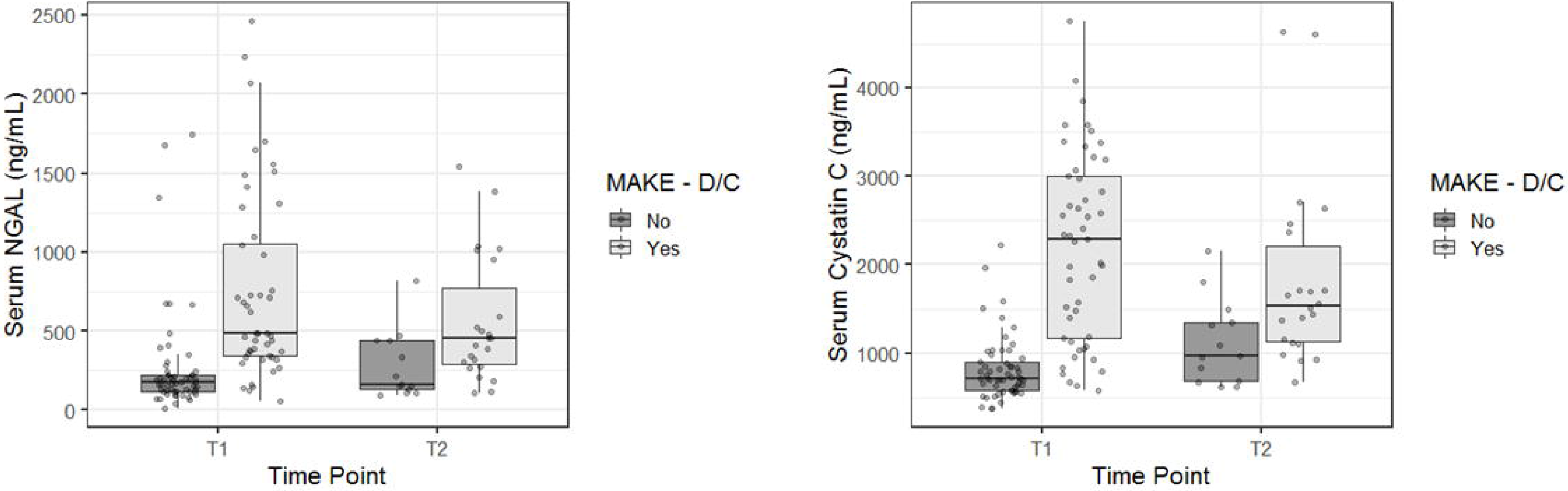
kidney Biomarker Trends from Time 1 (24-48 hours) to Time 2 (5-7days)

The simple clinical model for MAKE-DC including APACHE II and CCI produced an AUROC of 0.827, with both APACHE II (Odds Ratio (OR) 1.13; 95% CI 1.06-1.21) and CCI (1.41; 95% CI 1.15-1.74) independent predictors of MAKE-DC. Both serum NGAL (AUROC = 0.873) and serum cystatin C (AUROC=0.941) when independently added to the simple clinical model improved the AUROC (p= 0.019 and 0.001, respectively) as shown in **Figure 2**. None of the urinary biomarkers improved upon the simple clinical model for the prediction of MAKE-DC.

**Figure 2.**
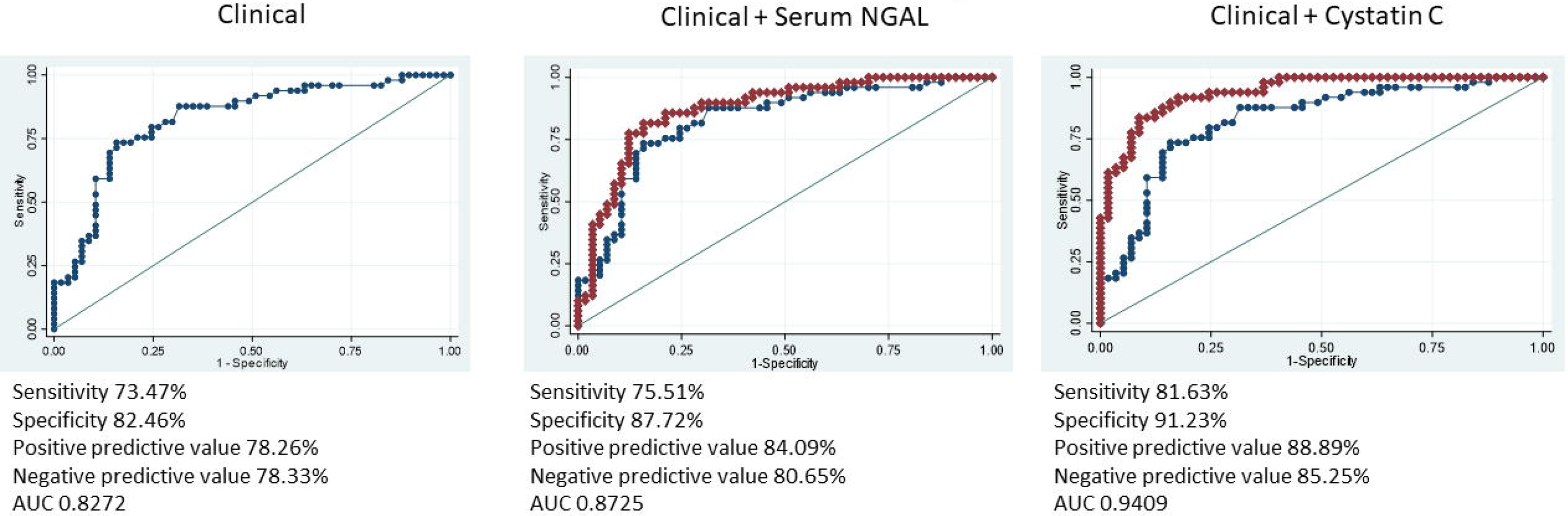
Receiver operating characteristic curves for MAKE-Discharge

In the exploratory analysis of biomarker trajectory from timepoint T1 to T2, only the change in serum NGAL (Δ NGAL) was significantly different in those patients experiencing MAKE-DC compared to those not. Those patients experiencing greater decreases in serum NGAL from time point T1 to T2 were less likely to experience MAKE-DC compared to those not (p=0.013) (**Table 4**). To account for baseline serum NGAL at time T1, both T1 and T2 serum NGAL values were assessed in a logistic regression model for MAKE-DC and the serum NGAL value at T2 remained significantly associated with MAKE-DC (OR 1.006; 95% CI 1.001-1.012).

**Table 4.**
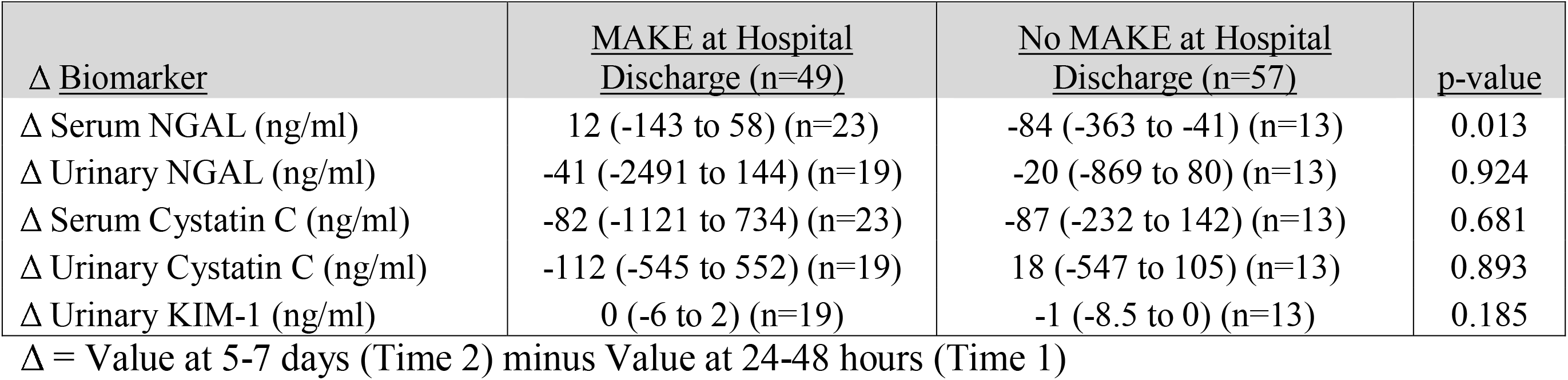
Change in Kidney Biomarkers and MAKE.

## Discussion

In this study of critically ill patients, serum and urinary biomarkers of AKI early in the course of AKI or absence of AKI in the ICU stay were significantly higher in those patients experiencing MAKE at several time periods, including hospital discharge, 90 days, 6 months, and 1 year. Of these, serum NGAL and cystatin C appeared to have the most utility when added to a simple clinical prediction model consisting of APACHE II and CCI. We also present evidence suggesting that trending these biomarkers in AKI may carry prognostic value. Overall, for the purpose of predicting MAKE events and trending biomarkers, serum NGAL and cystatin C appeared to perform strongest from the population in our study.

AKI biomarker panels, including multiple markers such as tubular injury and renal stress, have been theorized to have important implications for personalized medicine of AKI patients and the potential to improve prognostic, diagnostic, and therapeutic tools.^12^ Although not explored in our analysis, the combination of these biomarker panels has been shown in some settings to improve the predictive capability for AKI, particularly in the post-cardiac surgery setting.^13-16^ The value of biomarker trajectory modeling in critical illness is gaining traction, and although AKI subphenotypes based on serum creatinine have been examined, the value of repeated kidney biomarker assessments remains an area of active investigation.^17^ The repeated assessment of kidney biomarkers not only applies to the acute care setting, but to outpatient follow up as well. Follow up with a nephrologist after severe AKI has been associated with reduced all-cause mortality, and these biomarker assessments in the AKI recovery period may help to guide prognosis and therapy to reduce the development of chronic kidney disease post-AKI.^18^ Our sample size from the post-AKI visit at 4-6 weeks following AKI was unfortunately not large enough to evaluate this utility further. We did note that serum NGAL and cystatin C values at the T3 time point in AKI patients tended to be numerically higher than values obtained from the control group at T1 and T2 and are worthy of additional study in the post-AKI recovery setting.

This is certainly not the first study to investigate serum NGAL or cystatin C. However, the vast majority of the literature on these biomarkers is related to the prediction of AKI, for which they have generally not performed as well as anticipated.^1^ Given the increasing use of MAKE-DC or MAKE-30 as a primary outcome of clinical trials of AKI, particularly in the ICU setting, the ability to apply a biomarker to clinical prediction has great benefit in terms of prognostic enrichment and risk stratification in clinical trials. The association of biomarker levels with important patient-centered outcomes, including death, need of RRT, or reduced eGFR, out to one year is also important in the consideration of family discussions, discharge planning, and goals of care in the ICU setting.

A major strength of our study is the prospective design and follow-up of biomarkers and clinical outcomes at repeated time intervals, including out to one year. Although biomarkers are generally recognized to respond differently depending on the specific kidney insult (i.e. sepsis, ischemia, etc), our study encompassed medical, surgical, and cardiac ICUs which increases its generalizability to several different areas of practice and research. While each type of kidney insult may produce its own specific pattern of kidney biomarker expression, often the kidney injury in the ICU is multifactorial thus biomarkers applicable to a wide variety of clinical settings are desirable. We used well-established criteria for the diagnosis of AKI and the outcome of interest (MAKE).^5, 8^

Our findings are not without their noted limitations. First, the cohort was from a single center and relatively small sample size. Due to death or loss to follow-up, missing data were more prevalent than preferred for both biomarker measurement at time T2 or T3 as well as MAKE events after hospital discharge, which increases the chance of bias when analyzed using complete-case analysis as we have done. These factors limited the number of covariates we could include in the clinical prediction model, and the possibility exists that a more detailed clinical prediction model for MAKE events may not benefit from the addition of biomarker measurements. While we noted the broad population of the cohort as a strength, it could potentially represent a limitation if it diluted the effect of any one biomarker in a specific patient population, such as sepsis. Finally, we only included AKI stage 2 and greater without chronic kidney disease at baseline, thus patients with AKI stage 1 or chronic kidney disease may not be adequately addressed in this study. Future research offers the opportunity to evaluate the predictive performance of these biomarkers, particularly serum NGAL and cystatin C, and more specifically their trajectories, in larger numbers of critically ill patients with AKI to further investigate the dose-response relationship and trajectory relationships between biomarker elevation and pertinent clinical outcomes.

## Conclusion

Early measurement of kidney biomarkers in critically ill patients associates with MAKE at hospital discharge. This relationship appears to be greatest with serum NGAL and cystatin C, which display additive performance to a simple clinical prediction model. Trending serum NGAL may also help predict MAKE at discharge.

## Disclosure

The authors have disclosed that they do not have any potential conflicts of interest.

## Data Availability

The data that support the findings of this study are available from the corresponding author upon reasonable request

## Acknowledgments

N/A

